# Early, robust mucosal secretory IgA but not IgG response to SARS-CoV-2 spike in oral fluid is associated with faster viral clearance and COVID-19 symptom resolution

**DOI:** 10.1101/2024.02.21.24303146

**Authors:** Nora Pisanic, Annukka A. R. Antar, Marissa K. Hetrich, Zoe O. Demko, Xueyan Zhang, Kristoffer Spicer, Kate L. Kruczynski, Barbara Detrick, William Clarke, Maria Deloria Knoll, David L. Thomas, Fatimah S. Dawood, Vic Veguilla, Ruth A. Karron, Yukari C. Manabe, Christopher D. Heaney

## Abstract

High priority efforts are under way to support the development of novel mucosal COVID-19 vaccines, such as the US Government’s Project *NextGen* and the Center for Epidemic Preparedness Innovations’ (CEPI) goal to respond to the next pandemic with a new vaccine in 100 days. However, there is limited consensus about the complementary role of mucosal immunity in disease progression and how the immunogenicity of mucosal vaccines will be evaluated. This study investigated the role of oral mucosal antibody responses in viral clearance and in COVID-19 symptom duration.

Participants with PCR-confirmed SARS-CoV-2 infection provided oral fluid for testing with SARS-CoV-2 antibody multiplex assays, nasal swabs for RT-PCR and symptom information at up to eight follow-ups from April 2020 to February 2022.

High and moderate oral fluid anti-spike (S) SIgA post infection was associated with significantly higher likelihood of viral clearance and of COVID-19 symptom resolution across age groups. Those with high and moderate anti-S SIgA cleared the virus and recovered 14 days (95% CI: 10-18 days) and 9-10 days (95% CI: 6-14 days) earlier, respectively. Delayed but higher oral fluid anti-S IgG was associated with significantly longer time to viral clearance and recovery. The effect size of moderate or high SIgA was equivalent to prior COVID-19 vaccine immunity, which was also associated with faster clearance and recovery. Unvaccinated adults with prolonged COVID-19 symptoms had significantly lower anti-RBD SIgA 15-30 days after infection onset (*p*<0.001).

Robust mucosal SIgA early post infection appears to support faster clearance of SARS-CoV-2 and recovery from COVID-19 symptoms. This research underscores the importance of harmonizing mucosal immune response assays to evaluate new vaccines that can boost local mucosal immunity.

**Disclaimer:** The findings and conclusions in this report are those of the authors and do not necessarily represent the official position of the Centers for Disease Control and Prevention.

## Introduction

While COVID-19 vaccines have saved millions of lives in the US, they have not stopped SARS-CoV-2 breakthrough infections or transmission of ever evolving variants. Despite triple vaccination, multiple studies have shown that breakthrough SARS-CoV-2 infections are common with the rapidly evolving omicron variants.(1,2) Currently approved intramuscular COVID-19 vaccines elicit strong serum neutralizing antibody (nAb) titers and binding IgG to SARS-CoV-2 spike protein (S), which have been identified as correlates of protection from symptomatic and severe COVID-19, however, they confer little mucosal immunity at the site of virus entry.(3) Blood nAb titers and anti-S IgG levels decline within months, resulting in the need for multiple intramuscular boosters with updated composition to match circulating variants. To break this continuing infection and transmission cycle, novel mucosal vaccines are being developed with support of national and international programs, such as the US government’s Project NextGen and the Center for Epidemic Preparedness Innovations (CEPI) with the goal to respond to the next pandemic with a new vaccine in 100 days.(4)

Mucosal SARS-CoV-2 anti-S IgA was observed to be a correlate for prevention of omicron infection as determined by positive polymerase chain reaction (PCR) results for viral RNA in triple-vaccinated health care workers. (5) Early in the COVID-19 pandemic, higher anti-RBD antibody levels in nasal mucosal lining fluid were shown to be associated with lower viral loads after PCR-confirmed SARS-CoV-2 infection.(6) Another recent study showed that time to mounting an anti-S secretory IgA response in the nasal cavity correlated significantly with the duration of viral shedding determined by virus culture. However, in that study, time to seroconversion and duration of viral shedding were both estimated from models based on different study populations. (7) And less is known about the role of mucosal antibody responses in oral fluid as a correlate of protection from other aspects of COVID-19 including time to viral clearance (first PCR negative test after COVID-19 infection), time to symptom resolution, and risk of developing Post COVID Conditions.(8) We sought to determine whether mucosal humoral antibody responses to SARS-CoV-2 were associated with viral clearance and symptom resolution. A better understanding of the role of mucosal immune responses will help inform intranasal COVID-19 vaccination strategies that boost mucosal immunity moving forward and assay standardization and harmonization efforts to assess immune responses in different mucosal compartments.(9,10)

## Methods

### Study design

This study assessed oral mucosal SARS-CoV-2 antibody responses in individuals with PCR-confirmed SARS-CoV-2 infection who participated in one of two prospective cohort studies, OutSMART (11,12) or SEARCh (13,14). The OutSMART study was approved by the Johns Hopkins University School of Medicine Institutional Review Board (IRB; IRB00259948). The SEARCh study was approved by the IRB of Johns Hopkins University Bloomberg School of Public Health (IRB00014200); the IRB of the Centers for Disease Control and Prevention approved the study based on the review of the Johns Hopkins University Bloomberg School of Public Health IRB.

Briefly, OutSMART enrolled predominantly adult participants who were identified through the Johns Hopkins Outpatient SARS-CoV-2 testing network and received a positive SARS-CoV-2 RT-PCR result. Participants self-collected swabs for SARS-CoV-2 RT-PCR testing and oral fluid for antibody testing and provided symptom information on study days 0, 3, 5, 7, 10, 14, 21 and 28. OutSMART samples collected between April 23, 2020, and February 22, 2022, contributed to this analysis. The SEARCh study enrolled households with at least one child <5 years old to participate in surveillance for incident SARS-CoV-2 infection for up to 8 months. Participants provided weekly nasal swabs for PCR testing and symptoms information and, upon PCR-confirmed SARS-CoV-2 infection, up to eight weekly oral fluid samples. SEARCh specimens collected between December 16, 2020, and October 6, 2021, contributed to this analysis. More detailed information about the study designs has been published previously and is provided in the **Supplement**. (11–14)

### Oral fluid collection and multiplex SARS-CoV-2 antibody assays

Oral fluid (Oracol+, Malvern Medical Developments, UK) and nasal swabs for PCR testing were participant-collected and mailed to the testing lab. Oral fluid collected 0 to 125 days after infection onset was tested for SARS-CoV-2 receptor binding domain (RBD)- and Spike (S)-specific IgG, IgA and secretory-component (SC)-containing Ig responses using magnetic bead-based multiplex immunoassays optimized for oral fluid, as described previously.(15–17) Mucosal SC of polymeric Ig was detected using a mouse monoclonal anti-SC antibody, followed by labeled anti-mouse antibody, hereafter referred to as secretory IgA (SIgA) since IgA is the predominant secretory Ig in oral fluid.(18) Qualitative thresholds for oral fluid anti-S and RBD IgG had been established previously.(16,19) Assay performance for detecting anti-S-RBD IgG was 99.4% sensitivity (481/484 correct positives) and 99.3% specificity (995/1002 correct negatives) using oral fluid from presumably naïve individuals (∼1,000 samples), oral fluid collected >14 days after completing a COVID-19 vaccination series (∼175 samples) and/or >14 days after symptoms onset or after the first SARS-CoV-2 RT-PCR positive test (∼325 samples).

### Statistical analysis

#### Time to viral clearance

SARS-CoV-2 infection onset was defined as participant-reported COVID-19 symptom start date or the first PCR positive date, whichever occurred earlier. If asymptomatic, the first PCR positive date was used. The date of viral clearance was defined as the median date between the last SARS-CoV-2 RT-PCR positive and first negative nasal swab if less than 20 days apart.(12) If no first negative nasal swab PCR test was available within 20 days after the last positive PCR test, the individual’s oral fluid samples were excluded from time to clearance analyses. Time to viral clearance in days was calculated as the difference between infection onset and date of clearance as defined above.

#### Symptom duration

Similarly, symptom duration was defined as the difference in days between infection onset and the median date between the last day on which any COVID-19 symptoms were reported and the first asymptomatic day or, when available, the participant-reported date of symptoms resolution.

#### COVID-19 symptoms

Both studies assessed symptoms, however, symptom severity was only assessed in OutSMART. COVID-19 symptoms considered here and assessed in both studies included fever and chills, sore throat, loss of smell or taste, cough, congestion (nose, sinus pressure), body or muscle aches, chest congestion or pain, shortness of breath or difficulty breathing and diarrhea. The number of reported COVID-19 symptoms reported was calculated for each participant. In OutSMART the number of symptoms of the study visit during which the most COVID-19 symptoms were reported was used. Most participants reported the highest number of COVID-19 symptoms at enrollment through study day 3 (143/204 participants; 71%). SEARCh inquired weekly about onset of COVID-19 and other symptoms. To capture COVID-19 symptoms reported within an equivalent time frame, COVID-19 symptoms reported one week prior and up to two weeks after the first PCR positive test counted towards the number of COVID-19 symptoms reported.

#### Infecting SARS-CoV-2 variant

If the infecting variant was unknown, the most prevalent variant (20) circulating at the time of PCR-confirmed SARS-CoV-2 infection was imputed with wild type imputed for infections occurring through February 2021; Alpha from March 2021 instead of April (20) to match the sequencing-derived variant results available (SEARCh) through June 2021; Delta from July through November 2021; and Omicron starting in December 2021 until study end. This approach resulted in matching sequencing-based and imputed SARS-CoV-2 variant strains for all sequencing-confirmed SARS-CoV-2 isolates (24/24 isolates in SEARCh and 7/7 confirmed Omicron isolates in OutSMART).

#### Statistical models

Cox proportional hazard models and Kaplan-Meier survival plots were used to assess associations between temporal dynamics of oral fluid SARS-CoV-2 antibody levels and other factors with time to viral clearance and COVID-19 symptom resolution. Asymptomatic individuals were assigned a symptom duration of 0 days and included in analyses (coded in time to event analyses as 1 = resolution of symptoms reached). Oral fluid anti-S antibody responses were classified into tertiles (low, moderate, and high), allowing for stratified visualization with Kaplan-Meier survival plots and easier comparison with other factors such as prior COVID-19 vaccine immunity. In another approach, participants were classified into tertiles based on their median, rather than time-varying, anti-S response post infection (low, moderate, high SIgA responder), while maintaining anti-S IgG as a time-varying covariate in the model. A third approach used time-stratified analysis to test whether the direction and magnitude of associations remained consistent over time.

In Cox proportional hazard models and in Kaplan-Meier survival analyses the event (viral clearance or symptom resolution) was coded as 1. Therefore, in contrast to survival analyses where the event is disease or death coded as 1, in the models presented here hazard ratios (HR)>1 indicate higher likelihood of viral clearance or symptom resolution and shorter time to clearance or symptom resolution in Kaplan-Meier analyses; HRs<0 indicate lower likelihood of and shorter time to clearance or symptom resolution. Linear regression models with generalized estimating equations (GEE) were also used to estimate the effect of anti-S and anti-RBD SIgA, IgA and IgG responses on time to clearance and symptom duration.

Models were adjusted for COVID-19 vaccination status (received at least 1 COVID-19 vaccine dose versus unvaccinated at time of infection), infecting variant, age group (<5, 5-17, >=18, >65 years), sex, race, and ethnicity. GEE models were also adjusted for time since infection onset. All models included clustering equations to account for non-independence of repeated oral fluid measurements within individual. Collinearity between independent covariates was estimated by calculating variance inflation factors for all models. Only models with variance inflation factors under 4 are presented here. Analyses were performed in R version 4.2.1 using packages tidyverse, survival, survminer, forestploter, adjustedCurves, ggpubr, and car.

## Results

Oral fluid samples (n=1,256) from 257 adults and children with PCR-confirmed COVID-19 infection who participated either in the OutSMART or SEARCh cohort studies were analyzed for anti-RBD and anti-S secretory component-containing Ig (predominantly secretory IgA [SIgA]), IgA and IgG responses. Almost all participants experienced mild or moderate COVID-19 disease; children were more often asymptomatic, as described in more detail elsewhere.(11–13) Participant characteristics stratified by study cohort and combined are shown in **Table 1**. The median time from infection onset to viral clearance, as defined above, was 15 days (interquartile range [IQR] = [9.5; 21.5]) and median time to resolution of COVID-19 symptoms was 18 days (IQR = [9.25; 37.0]).

**Table 1.** Participant characteristics by study cohort.

| Participant characteristic | OutSMART | SEARCH | Combined |
| --- | --- | --- | --- |
| Participants, n | 208 | 49 | 257 |
| Prior Immunity |  |  |  |
| Vaccinated against COVID-19, n (%) | 38 (18%) | 4 (8%) | 42 (16%) |
| Not COVID-19 vaccinated, n (%) | 170 (82%) | 45 (92%) | 215 (84%) |
| Sex |  |  |  |
| Female | 124 (60%) | 23 (47%) | 147 (57%) |
| Male | 84 (40%) | 26 (53%) | 110 (43%) |
| Age Group |  |  |  |
| 0-4 years, n (%) | 3 (1%) | 19 (39%) | 22 (9%) |
| 5-17 years, n (%) | 7 (3%) | 11 (22%) | 18 (7%) |
| >=18 years, n (%) | 167 (80%) | 19 (39%) | 186 (72%) |
| >65 years, n (%) | 31 (15%) | 0 (0%) | 31 (12%) |
| Race / Ethnicity |  |  |  |
| White, non-Hispanic | 94 (45%) | 35 (71%) | 129 (50%) |
| Black, non-Hispanic | 74 (36%) | 1 (2%) | 75 (29%) |
| Multiracial, non-Hispanic | 12 (6%) | 9 (18%) | 21 (8%) |
| Hispanic | 28 (13%) | 1 (2%) | 29 (11%) |
| SARS-CoV-2 Variant |  |  |  |
| Wildtype | 136 (65%) | 18 (37%) | 154 (60%) |
| Alpha | 41 (20%) | 18 (37%) | 59 (23%) |
| Delta | 21 (10%) | 13 (26%) | 34 (13%) |
| Omicron | 10 (5%) | 0 (0%) | 10 (4%) |
| Days to viral clearance, median (IQR) | 15.5 (12 - 23.5) | 10.5 (7.5-16.5) | 15 (9.5 - 21.5) |
| Days to COVID-19 symptoms resolution, median (IQR) | 25 (13.5 - 25) | 5 (0 - 10) | 18 (9.25-37) |
| Days to first oral fluid collection after symptoms onset or first SARS-CoV-2 positive PCR if asymptomatic, median (IQR) | 9 (5 – 16.5) | 7 (5 – 9) | 8 (5 – 13) |
| Days to oral fluid collection, median (IQR)* | 18 (11 – 34) | 33.5 (15 – 55) | 22 (12 – 43) |
| Oral fluid samples, n | 838 | 418 | 1256 |

**Note:** IQR = interquartile range (25%ile – 75%ile). \*Oral fluid was collected weekly (SEARCH) after SARS- CoV-2 PCR positive test or on study days 0, 3, 5, 7, 10, 14, 21, 28 (OutSMART).

### Factors associated with SARS-CoV-2 clearance and COVID-19 symptom resolution

Higher anti-S SIgA level and COVID-19 vaccination status were associated with greater likelihood of viral clearance and resolution of symptoms (**Figure 1**). Moderate compared to low anti-S SIgA was associated with a 1.8-fold greater likelihood of viral clearance (adjusted hazard ratio [aHR] with 95% confidence interval [CI] = 1.8 [1.2-2.5]; *p*=0.002) and 1.7-fold greater likelihood of COVID-19 symptoms resolution (aHR [95% CI] = 1.7 [1.2-2.3], *p*=0.003). High compared to low anti-S SIgA was associated with a 2.2-fold and 2.1-fold greater likelihood of viral clearance (aHR [95% CI] = 2.2 [1.5-3.1], *p*<0.001) and symptom resolution (aHR [95% CI] = 2.1 [1.5-2.9], *p*<0.001), respectively. Prior immunity from COVID-19 vaccination was also associated with greater likelihood of clearance (aHR [95% CI] = 2.5 [1.8-3.4], *p*<0.001) and resolution of symptoms (aHR [95% CI] = 1.9 (1.3-2.7], *p*<0.001). In contrast, higher anti-S IgG was associated with reduced likelihood of viral clearance and symptoms resolution (**Figure 1)**.

**Figure 1.**
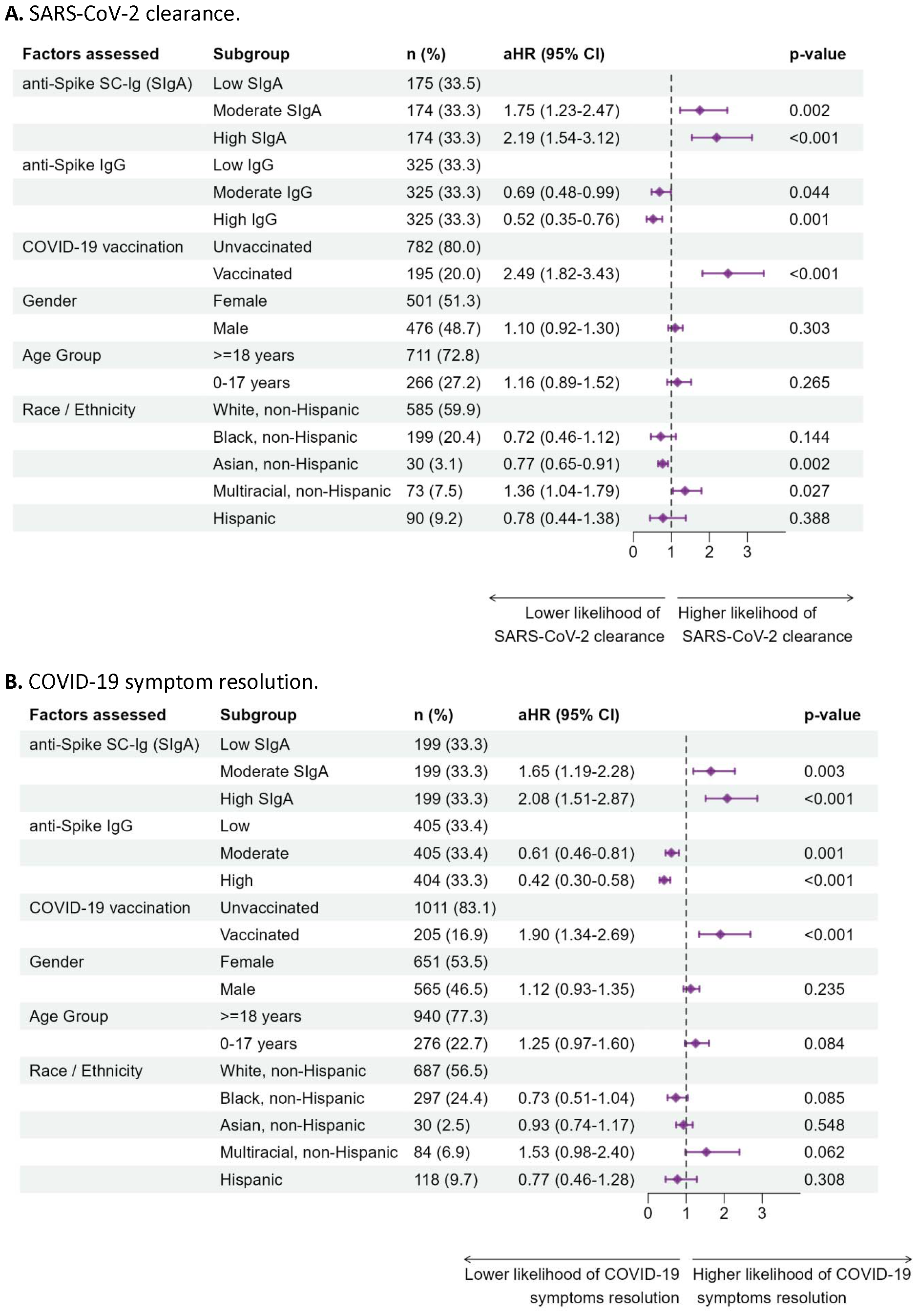
Cox proportional hazard model to assess factors associated with **A**. SARS-CoV-2 clearance (first SARS-CoV-2 PCR negative test) and **B**. COVID-19 symptom resolution. **Note:** aHR = hazard ratio estimated from Cox proportional hazard model adjusted for all covariates presented in the table and clustered at the individual level. CI = Confidence interval. SC = Secretory component. In time to event analyses 1 = time individual (**A**) clears the SARS-CoV-2 virus (tests SARS-CoV-2 PCR negative), or (**B**) reports being asymptomatic; 0 = individual continues to (**A**) test SARS-CoV-2 PCR positive, or (**B**) report COVID-19 symptoms. HRs > 1.0 indicate higher likelihood and HRs < 1.0 indicate lower likelihood of (**A**) SARS-CoV-2 clearance, or (**B**) resolution of COVID-19 symptoms.

In children under 5 years of age, higher anti-S SIgA was also associated with increased likelihood of viral clearance (aHR [95% CI] for each log_10_ increase in anti-S SIgA = 1.4 [1.0-1.9], *p*<0.05) and of symptoms resolution (aHR [95% CI] for each log_10_ increase in anti-S SIgA = 1.4 [1.2-1.6] p<0.001). Higher anti-S IgG in children was associated with lower likelihood of viral clearance (aHR [95% CI] for each log_10_ increase in anti-S IgG = 0.6 [0.4-0.9], *p*<0.01; **Tables S1 and S7**). These trends remained consistent and statistically significant in independent analyses of both study cohorts (**Tables S2-S3**) and of unvaccinated adults (**Table S8**). In the OutSMART cohort, age >65 years was a risk factor for prolonged PCR positivity (aHR [95% CI] = 0.6 [0.3-0.9], *p*<0.05; **Table S3**); the SEARCh cohort did not enroll participants over 65 years. In the SEARCh cohort, younger age (children 0-17 years old) was associated with greater likelihood of viral clearance (aHR [95% CI] = 1.35 [1.02-1.80], *p*<0.05; **Table S4**). Adjusting for the infecting variant did not affect the magnitude or direction of these associations significantly but showed that Delta and Omicron infections were associated with significantly shorter time to viral clearance than wildtype infections (**Table S5**).

### Differences in time to clearance and symptoms resolution

In Kaplan-Meier survival estimates stratified by anti-S SIgA tertile, moderate oral fluid anti-S SIgA compared to low SIgA was associated with clearing the virus 10 days earlier (95% CI: 7-14 days) and with becoming asymptomatic 9 days (95% CI: 6-14 days) earlier. High compared to low anti-S SIgA was associated with clearing the virus 14 days (95% CI: 10-18 days) earlier and with becoming asymptomatic 14 days (95%CI: 10-17 days) earlier (**Figure 2**). The direction and effect size of these associations remained consistent over time as confirmed with time since infection restricted subsets of oral fluid and using participants’ median rather than time varying SIgA response post infection (see **Supplement Time-restricted models and Median SIgA response post infection**).

**Figure 2.**
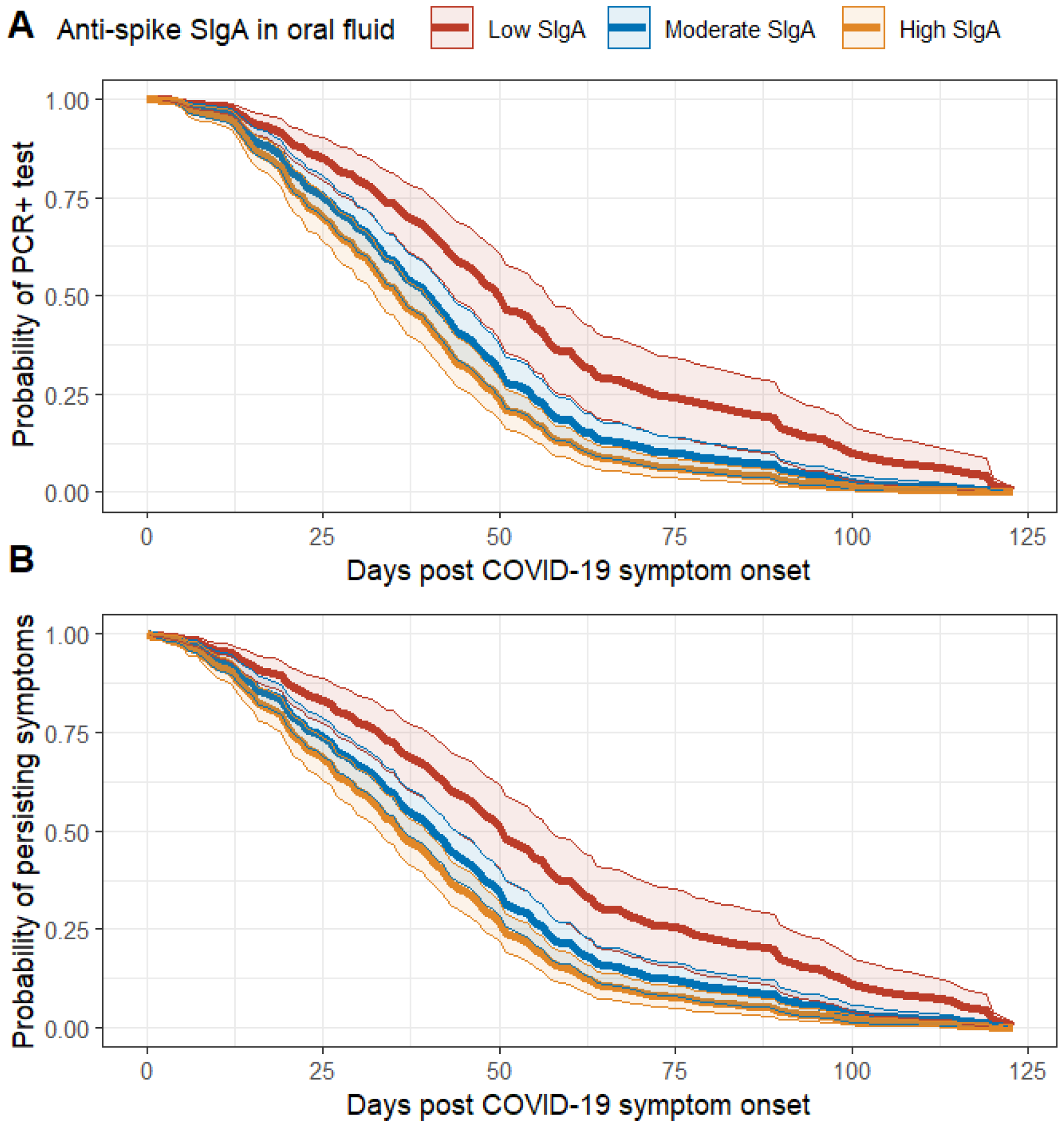
Kaplan-Meier survival curves stratified by oral fluid anti-spike SIgA response post COVID-19 symptoms onset (low; moderate and high SIgA tertile). **A**. Probability of testing SARS-CoV-2 PCR positive over time in days. **B**. Probability of persisting COVID-19 symptoms over time in days.

### Association of oral fluid anti-RBD Ig with time to clearance and COVID-19 symptoms duration

Mucosal SARS-CoV-2 RBD-specific SIgA and IgA in children and adults were higher among the two-thirds of study participants (67%) who cleared the virus within 18 days post infection onset compared to those who tested PCR positive longer (**Figure S2**). Oral fluid SARS-CoV-2 IgG levels, which correlate with serum SARS-CoV-2 IgG levels (16,17), were similar in individuals who cleared the virus in ≤18 days and >18 days among adults. The number of children that did not clear the virus within 18 days (n=4) was too small for representative comparison of the two groups (**Figure S2**). Adults who resolved symptoms within four weeks had significantly higher anti-RBD SIgA and IgA, but not IgG, 15-30 days after infection onset than individuals who remained symptomatic longer than four weeks (*p*<0.0001; **Figure S3)**.

### Association between delayed seroconversion and viral clearance

Even though higher anti-S IgG in oral fluid was associated with longer time to viral clearance, shorter time to a first positive oral fluid anti-S IgG result (shorter time to seroconversion) remained associated with faster viral clearance (**Table S6**), as shown previously in a subset of the OutSMART cohort.(12) GEE models estimated a 0.8-day increase (beta [SE] = 0.8 [0.2], *p*<0.001) in time to clearance for each one-day delay in time to anti-S IgG seroconversion.

### Linear regression model to estimate associations between oral fluid anti-S and anti-RBD Ig and time to SARS-CoV-2 clearance or COVID-19 symptoms duration

Each log_10_ unit increase in anti-S and anti-RBD SIgA and IgA was associated with three to five days faster viral clearance and COVID-19 symptoms resolution. Every log_10_ increase of oral fluid anti-RBD (Sino Biological, China) SIgA was associated with beta [SE] -2.7 days [1.3], *p*<0.05 faster viral clearance; while every log_10_ increase in anti-S (Mt. Sinai) IgA was associated with 3.4 days faster viral clearance (beta [SE]: -3.4 days [1.3]; *p*<0.01, **Table S9**). Similarly, each log_10_ increase in oral fluid anti-S-RBD was associated with 3.4 to 4.9 days earlier resolution of COVID-19 symptoms (beta [SE]: -3.4 [1.6]; *p*<0.05 to -4.9 [1.9] days, *p*<0.01, **Table S10**). Higher anti-S IgG was again associated with longer time to clearance and to becoming asymptomatic. Children cleared the virus about one week earlier than adults (beta [SE] = -6.8 days (2.7); *p*<0.01) and recovered significantly faster from COVID-19 symptoms than adults (beta [SE] = -16.5 (2.7); *p*<0.001), while among adults self-reported Black race appeared associated with longer time to clearance and symptoms duration (**Table S10**).

Lastly, we investigated whether the mucosal immune response to SARS-CoV-2 differed depending on the maximum number of COVID-19 symptoms reported (0-3, 4-6, or >6 symptoms). Participants who reported experiencing >6 compared to 4-6 and 0-3 COVID-19 symptoms had higher anti-RBD IgG 14-30 days after infection onset. This trend was consistent among children (Kruskal-Wallis *p*<0.05) and among adults (*p*<0.01) but anti-RBD IgG <14 days post infection and anti-RBD SIgA or IgA did not appear associated with the number of COVDID-19 symptoms reported (**Figure S4)**.

## Discussion

Among children and adults, early and robust mucosal SARS-CoV-2 SIgA and IgA responses in oral fluid were associated with faster viral clearance and earlier recovery from COVID-19 symptoms. Early anti-S IgG seroconversion was associated with faster viral clearance, as shown before in the OutSMART cohort (12), while higher anti-S IgG was associated with longer time to clearance and to resolution of COVID-19 symptoms. These findings are consistent with prior research showing positive correlations between blood anti-S IgG and nAb titers in convalescence and COVID-19 disease severity and symptoms duration (21–23), and between delayed nAb and anti-S IgG responses and severe outcomes in children and fatal outcomes COVID-19 in adults.(23,24) Our group has also demonstrated that prolonged time to viral clearance during the acute phase of SARS-CoV-2 infection is associated with long COVID.(25) We showed that COVID-19 vaccine-induced immunity at the time of infection was associated with faster clearance and earlier resolution of symptoms with equivalent effect size as moderate and high SIgA. Early, robust anti-S SIgA responses in oral fluid may thus be a correlate for protection from prolonged viral persistence and from prolonged duration of COVID-19 symptom with comparable magnitude as conferred by intramuscular COVID-19 vaccination.

These findings are consistent with a model in which early and robust local mucosal anti-S SIgA and timely IgG seroconversion contribute to early viral clearance, e.g., by blocking viral attachment to mucosal cell surfaces (SARS-CoV-2 binding to ACE2) and thus preventing infection of target epithelial cells and later spread to other sites of infection. In this model, individuals who cannot mount early and robust local mucosal anti-S SIgA responses and/or who experience prolonged time to rise of anti-S IgG are more likely to experience prolonged unmitigated infection, higher virion burden at sites of infection, and/or systemic spread. These individuals would have a higher burden of viral antigen interacting with B cells throughout lymphatic tissue and therefore would be more likely to eventually mount higher titers of anti-SARS-CoV-2 IgG.

Limitations of this research included that the SEARCh and OutSMART study COVID-19 symptoms assessments were not administered at the same time intervals relative to COVID-19 infection and that symptom severity was only captured in one of the two cohorts, thus limiting the sample size to investigate associations between oral fluid antibody responses and COVID-19 severity. Only the number of SARS-CoV-2 symptoms, which were reported in both cohorts, and symptom duration but not symptom severity in relation to oral mucosal antibody responses were investigated here.

We showed that unvaccinated adults who experienced COVID-19 symptoms for more than four weeks had significantly lower levels of oral fluid anti-RBD SIgA and IgA than those who resolved symptoms earlier. However, we could not assess the role of SIgA in the development of long COVID here. Post COVID Conditions, including symptoms such as fatigue, post-exertional malaise, and brain fog that persist beyond or start after the four-week COVID-19 symptom duration period, were ascertained in only one of the study cohorts and were therefore excluded from this analysis.

Since in OutSMART PCR measurements were only available on select study days rather than weekly, approximately 50 out of 300 potential participants with either missing or delayed first negative PCR tests (≥20 days after the last PCR positive test) had to be excluded from time to event analyses. Similarly, SEARCh study participants who experienced COVID-19 infection near the end of the study period were excluded since date of first PCR negative nasal swab and of symptoms resolution were not ascertained and thus time to clearance and symptoms duration could not be calculated. Others modeled the viral shedding duration and the time to SIgA seroconversion in nasopharyngeal swabs based on the viral dynamics observed in other studies.(7) Here we chose to only include participants for which oral fluid antibody measurements were obtained and time to viral clearance (first SARS-CoV-2 PCR negative test) and symptoms duration could be calculated.

Finally, this study used qualitative FDA-EUA-approved SARS-CoV-2 PCR assays to infer time to viral clearance rather than SARS-CoV-2 virus culture, which was only available for a subset of PCR-positive swabs and originated from different laboratories. Therefore, the time to viral clearance used in this analysis represents the time from participant-reported symptoms onset or first PCR positive test to the first SARS-CoV-2 PCR negative test and is likely longer than the SARS-CoV-2 transmission window during which infectious virus is shed.

A Swedish study of COVID-19 vaccinated healthcare workers found that high post-vaccination SARS-CoV-2 specific SIgA levels may protect from SARS-CoV-2 breakthrough infections and recent research showed that the model-inferred duration of viral shedding duration was reduced when estimated nasal anti-S SIgA seroconversion occurred earlier.(5,7) Taken together, these studies and our research indicate that vaccine strategies that boost mucosal humoral immune responses, such as intranasal or inhaled SARS-CoV-2 vaccines known to boost S-RBD-binding local SIgA, may help protect from breakthrough SARS-CoV-2 infection, reduce the window for transmission, and reduce the duration and severity of COVID-19 symptoms following infection.(9,10,26) These findings underscore the importance of measures of mucosal immunogenicity, particularly standardization and harmonization as large-scale mucosal vaccination efforts such as Project NextGen proceed.

Future studies should investigate if mucosal anti-S SIgA could be a mechanistic (i.e., causal) or nonmechanistic correlate (i.e., associated, but not the primary mechanistic cause) (27,28) for faster viral clearance and resolution of COVID-19 symptoms. Mucosal secretory antibody responses are among several immune functions that could contribute to effective clearance of SARS-CoV-2 in addition to innate immune responses and other adaptive immune responses including antigen-specific killing of infected cells by CD8^+^ T cells.(29) Our work indicates a role for SIgA in clearance of virus and resolution of symptoms. Future work should focus on whether mucosal anti-S/RBD SIgA responses during acute SARS-CoV-2 infection are associated with later development of Post COVID Conditions, which include the persistence of COVID-19 symptoms, recurrence, or development of new symptoms four of more weeks after an initial SARS-CoV-2 infection.(30)

## Data Availability

All data produced in the present study are available upon reasonable request to the authors after consultation with and approval of all study PIs.

