## Supplemental Material for "Early, robust mucosal secretory IgA but not IgG response to SARS-CoV-2 spike in oral fluid is associated with faster viral clearance and COVID-19 symptom resolution"

**Supplement**

**Methods**

Mucosal anti-SARS-CoV-2 S and receptor binding domain (RBD) IgG, IgA, and secretory IgA (SIgA) responses were assessed in study participants with PCR-confirmed SARS-CoV-2 infection that were identified through the Johns Hopkins Outpatient SARS-CoV-2 testing network and consented to participate in the Outpatient SARS-CoV-2 Mild and Asymptomatic immune Response and Transmission study (OutSMART)(1,2) or participated in the SARS-CoV-2 Epidemiology and Response in Children (SEARCh) cohort study.(3) Both studies enrolled participants in Baltimore City and surrounding counties. The OutSMART study was approved by the Johns Hopkins University School of Medicine Institutional Review Board (IRB00259948). The SEARCh study was approved by the institutional review board of Johns Hopkins University Bloomberg School of Public Health (IRB00014200); the institutional review board of the Centers for Disease Control and Prevention approved the study based on the review of the Johns Hopkins University Bloomberg School of Public Health.

Study cohorts and design

OutSMART. (1,2) Eligibility criteria included having received a positive nasopharyngeal SARS-CoV-2 reverse-transcription quantitative polymerase chain reaction (RT-qPCR) result from the Johns Hopkins Medical Microbiology laboratory, age ≥18 years (amended later to allow participation of household members of any age), being able to receive study materials while remaining in isolation, and willingness to perform self-collection of specimens. OutSMART participants self-collected oral fluid, mid-turbinate nasal and oropharyngeal (nasal-OP) swabs; the latter two swabs were placed into 3 mL viral transport medium (VTM). Nasal-OP swabs were tested with the Abbott Molecular RealTime SARS-CoV-2 assay. (2) Study procedures involved collection of specimens at enrollment (study day 0) and at follow-up study days scheduled 3, 5, 7, 10, 14, 21 and 28 days after enrollment. COVID-19 symptoms (qualitative and severity) were assessed *via* questionnaires on each study day. OutSMART oral fluid and nasal-OP samples collected between April 23, 2020, and February 22, 2022, contributed to this analysis.

SEARCh. (3,4) The SEARCh study enrolled households with at least one child <5 years old to participate in surveillance for incident SARS-CoV-2 infection for up to eight months. COVID-19 symptoms were assessed weekly. If present, surveys were administered that assessed presence of respiratory and systemic symptoms that allowed for calculation of the number of COVID-19 symptoms during incident infection. The persistence of any symptoms (yes/no) was assessed *via* active surveillance until resolution. Participants provided weekly anterior nasal swabs that were mailed to the Marshfield Clinic Research Institute and tested by qualitative RT-PCR for presence of SARS-CoV-2. (5) Specimens with cycle threshold values lower than 30 were processed for viral whole genome sequencing at the Centers for Disease Control and Prevention and classified as wildtype, Alpha, or Delta lineages. Participants provided monthly oral fluid samples and up to eight weekly samples triggered by incident PCR-confirmed SARS-CoV-2 infection. SEARCh specimens collected between December 16, 2020, and October 6, 2021, contributed to this analysis.

Blood SARS-CoV-2 antibody measurements prior to the PCR-confirmed SARS-CoV-2 infection were available for a subset of participants and confirmed self-reported prior immunity or lack thereof. Blood antibody measurements for almost all participants were available for the convalescent phase and supported the PCR-confirmed SARS-CoV-2 infection classification (confirmed seroconversion). We previously showed that almost all OutSMART participants seroconverted after SARS-CoV-2 infection and that oral fluid SARS-CoV-2 IgG measurements were an excellent alternative to blood-based SARS-CoV-2 serological assays.(6–8)

Oral fluid collection and multiplex SARS-CoV-2 antibody assays

Both studies provided sample collection kits containing oral and nasal swabs and written instructions for self-collection in person at enrollment or by mail. Study coordinators provided instructions on sample self-collection, packaging, and shipment by video call. Oral fluid was self-collected in both studies with an Oracol+ saliva collection swab (Malvern Medical Developments, UK) by rubbing the tooth-gum line for 1-2 minutes and then shipped back to the testing laboratory with ice packs.

**Supplementary Results**

Time-restricted models

*SARS-CoV-2 clearance.* Restricting the analysis to oral fluid samples collected within 30 days of infection onset (and thus the sample size), moderate compared to low anti-S SIgA was not associated with significantly higher likelihood of viral clearance (aHR [95% CI = 1.6 [0.84-3.0], *p*=0.17) but high vs. low anti-S SIgA remained significant (aHR [95% CI = 1.9 [1.0-3.3], *p*<0.05). Similar magnitudes of association were seen when restricting the analysis to oral fluid collected 14-30 days post infection onset (moderate vs. low SIgA: aHR [95% CI = 1.9 [1.1-3.3], *p*=0.02; high vs. low SIgA: aHR [95% CI = 1.7 [1.0-3.1], *p*=0.06). After 30 days post infection onset, moderate and high compared to low oral fluid anti-S SIgA was associated with 1.9-fold and 2.5-fold greater likelihood of viral clearance, respectively (moderate vs. low: aHR [95% CI] = 1.9 [1.3-2.7], *p*<0.001); high vs. low: aHR [95% CI] = 2.5 [1.7-3.6], *p*<0.001), confirming that the estimated associations were similar in magnitude over time.

*Symptoms resolution.* Restricting the analysis to oral fluid collected within 30 days post infection onset, 14-30 days, or >30 days post infection onset resulted in associations of similar magnitude and direction, e.g., at 14-30 days post infection: moderate vs. low anti-S SIgA aHR [95% CI] = 1.7 [1.1-2.7], *p*=0.02; high vs. low: aHR [95% CI] = 1.6 [0.9-2.8], *p*=0.09; in samples collected >30 days post infection: moderate vs. low anti-S SIgA: aHR [95% CI] = 1.9 [1.3-3.2], *p*<0.001; high vs. low: aHR [95% CI] = 2.4 [1.6-3.6], *p*<0.001.

Median SIgA response post infection

*Time to clearance.* When stratified by median rather than time-varying anti-SIgA response, participants with overall low anti-S SIgA in oral fluid following COVID-19 infection tested SARS-CoV-2 PCR positive for 9 days longer than those with overall moderate or high anti-S SIgA response (**Figure S1A**). However, in contrast to the time-varying SIgA analysis (**Figure 2A**), there was little difference in time to viral clearance between participants with moderate compared to high anti-S SIgA response (**Figure S1A**).

*Symptoms Resolution.* Overall high anti-S SIgA compared to low levels (3^rd^ tertile vs. 1^st^ tertile of within-person median SIgA response post infection) were associated with resolution of COVID-19 symptoms 13 days earlier and overall moderate compared to low SIgA levels (2^nd^ vs. 1^st^ tertile) were associated with recovering 8 days earlier from COVID-19 symptoms (**Figure S1.B)**.

**Table S1.** Cox proportional hazard model to assess risk factors associated with time to SARS-CoV-2 clearance in children under 5 years of age (SEARCh cohort).

| **Risk Factors** | **Mean (SD)** | **HR (95% CI, *p*-value)** | **aHR (95% CI, *p*-value)** |
| --- | --- | --- | --- |
| Anti-Spike SIgA (SC-Ig)* | 1.1 (1.1) | 1.16 (0.98-1.38, p<0.080) | 1.37 (1.02-1.86, **p<0.05**) |
| Anti-Spike IgG* | 2.6 (1.0) | 0.91 (0.74-1.12, p<0.365) | 0.57 (0.37-0.86, **p<0.01**) |
| **Sex** | **n (%)** |  |  |
| Female | 65 (40.4) | (Reference) | (Reference) |
| Male | 96 (59.6) | 1.01 (0.72-1.41, p<0.964) | 0.88 (0.49-1.58, p<0.67) |
| **Race / Ethnicity** | **n (%)** |  |  |
| White, non-Hispanic | 118 (73.3) | (Reference) | (Reference) |
| Asian, non-Hispanic | 10 (6.2) | 1.07 (0.56-2.05, p<0.845) | 1.02 (0.48-2.18, p<0.95) |
| Multiracial, non-Hispanic | 23 (14.3) | 2.22 (1.33-3.71, p<0.002) | 2.05 (1.05-4.03, p<0.05) |
| Hispanic | 10 (6.2) | 1.32 (0.64-2.72, p<0.457) | 0.85 (0.40-1.82, p<0.68) |

**Note.** aHR = hazard ratio estimated from Cox proportional hazard model adjusted for all covariates presented in the table and clustered at the level of the individual participant. *log_10_ median fluorescence intensity (MFI). SC = secretory component. HR > 1.0 indicates higher probability of (and shorter time to) and HR < 1.0 indicates lower probability of (and longer time to) SARS-CoV-2 clearance. CI=confidence interval.

**Table S2.** Cox proportional hazard model to assess risk factors associated with time to SARS-CoV-2 clearance in adults (>=18 years) participating in the SEARCh study.

| **Risk Factors** | **Mean (SD)** | **HR (95% CI, *p*-value)** | **aHR (95% CI, *p*-value)** |
| --- | --- | --- | --- |
| Anti-Spike SIgA (SC-Ig) ^a^ | 2.2 (1.0) | 1.19 (0.98-1.45, p<0.086) | 1.40 (1.19-1.64, **p<0.001**) |
| Anti-Spike IgG* | 3.4 (1.3) | 0.90 (0.73-1.10, p<0.298) | 0.62 (0.50-0.76, **p<0.001**) |
| **COVID-19 vaccination** | **n (%)** |  |  |
| Not vaccinated | 152 (88.9) | (Reference) | (Reference) |
| Vaccinated | 19 (11.1) | 2.43 (1.25-4.72, p<0.009) | 2.49 (1.69-3.67, **p<0.001**) |
| **Sex** | **n (%)** |  |  |
| Female | 87 (50.9) | (Reference) | (Reference) |
| Male | 84 (49.1) | 1.23 (0.87-1.74, p<0.243) | 1.24 (0.90-1.70, p<0.19) |
| **Race / Ethnicity** | **n (%)** |  |  |
| White, non-Hispanic | 141 (82.5) | (Reference) | (Reference) |
| Black, non-Hispanic | 4 (2.3) | 6.41 (1.51-27.28, p<0.012) | 3.49 (2.23-5.48, p<0.001) |
| Asian, non-Hispanic | 20 (11.7) | 0.78 (0.46-1.32, p<0.349) | 0.73 (0.52-1.02, p<0.07) |
| Multiracial, non-Hispanic | 6 (3.5) | 4.68 (1.10-19.85, p<0.036) | 0.00 (0.00-0.00, p<0.001) |
| Hispanic | 0 (0.0) | n/a | n/a |

**Note.** aHR = hazard ratio estimated from Cox proportional hazard model adjusted for all covariates presented in the table and clustered at the level of the individual participant. *log_10_ median fluorescence intensity (MFI). SC = secretory component. HR > 1.0 indicates higher probability of (and shorter time to) and HR < 1.0 indicates lower probability of (and longer time to) SARS-CoV-2 clearance. CI=confidence interval.

**Table S3.** Cox proportional hazard model to assess risk factors associated with time to SARS-CoV-2 clearance in adults (>=18 years) participating in the OutSMART study.

| **Risk Factors** | **Mean (SD)** | **HR (95% CI, *p*-value)** | **aHR (95% CI, *p*-value)** |
| --- | --- | --- | --- |
| Anti-Spike SIgA (SC-Ig) * | 1.7 (1.0) | 1.29 (1.07-1.55, p<0.007) | 1.29 (1.06-1.59, **p<0.05**) |
| Anti-Spike IgG* | 3.0 (1.2) | 0.89 (0.80-1.00, p<0.043) | 0.58 (0.43-0.77, **p<0.001**) |
| **COVID-19 vaccination** | **n (%)** |  |  |
| Not vaccinated | 407 (69.3) | (Reference) | (Reference) |
| Vaccinated | 180 (30.7) | 1.44 (1.13-1.84, p<0.003) | 2.81 (1.65-4.80, **p<0.001**) |
| **Sex** | **n (%)** |  |  |
| Female | 323 (54.9) | (Reference) | (Reference) |
| Male | 265 (45.1) | 0.82 (0.66-1.02, p<0.071) | 1.00 (0.66-1.51, p=1.00) |
| **Age** |  |  |  |
| Up to 65 years | 497 (84.5) | (Reference) | (Reference) |
| Over 65 years | 91 (15.5) | 0.62 (0.45-0.84, p<0.002) | 0.55 (0.32-0.92, **p<0.05**) |
| **Race / Ethnicity** | **n (%)** |  |  |
| White, non-Hispanic | 286 (48.6) | (Reference) | (Reference) |
| Black, non-Hispanic | 196 (33.3) | 0.77 (0.60-0.99, p<0.040) | 0.48 (0.29-0.80, **p<0.01**) |
| Asian, non-Hispanic | 0 (0.0) | n/a | n/a |
| Multiracial, non-Hispanic | 26 (4.4) | 0.80 (0.48-1.35, p<0.408) | 1.17 (0.52-2.63, p<0.710) |
| Hispanic | 80 (13.6) | 0.85 (0.63-1.15, p<0.294) | 0.45 (0.24-0.87, p<0.017) |

**Note.** aHR = hazard ratio estimated from Cox proportional hazard model adjusted for all covariates presented in the table and clustered at the level of the individual participant. *log_10_ median fluorescence intensity (MFI). SC = secretory component. HR > 1.0 indicates higher probability of (and shorter time to) and HR < 1.0 indicates lower probability of (and longer time to) SARS-CoV-2 clearance. CI=confidence interval.

**Table S4.** Cox proportional hazard model to assess risk factors associated with time to SARS-CoV-2 clearance among adults and children in the SEARCh cohort.

| **Risk Factors** | **Mean (SD)** | **HR (95% CI, *p*-value)** | **aHR (95% CI, *p*-value)** |
| --- | --- | --- | --- |
| Anti-Spike SIgA (SC-Ig) * | 1.5 (1.2) | 1.10 (0.99-1.21, p<0.075) | 1.46 (1.30-1.63, **p<0.001**) |
| Anti-Spike IgG* | 3.0 (1.2) | 0.89 (0.80-1.01, p<0.061) | 0.62 (0.50-0.77, **p<0.001**) |
| **COVID-19 vaccination** | **n (%)** |  |  |
| Not vaccinated | 390 (96.3) | (Reference) | (Reference) |
| Vaccinated | 15 (3.7) | 2.12 (1.12-4.01, p<0.020) | 3.16 (1.91-5.21, **p<0.001**) |
| **Sex** | **n (%)** |  |  |
| Female | 188 (46.4) | (Reference) | (Reference) |
| Male | 217 (53.6) | 0.99 (0.80-1.23, p<0.949) | 1.07 (0.85-1.36, p<0.554) |
| **Age Group** | **n (%)** |  |  |
| >=18 years | 176 (43.5) | (Reference) | (Reference) |
| 0-17 years | 229 (56.5) | 1.15 (0.93-1.42, p<0.195) | 1.35 (1.02-1.80, **p<0.05**) |

**Note.** aHR = hazard ratio estimated from Cox proportional hazard model adjusted for all covariates presented in the table and clustered at the level of the individual participant. *log_10_ median fluorescence intensity (MFI). SC = secretory component. HR > 1.0 indicates higher probability of (and shorter time to) and HR < 1.0 indicates lower probability of (and longer time to) SARS-CoV-2 clearance. CI=confidence interval.

**Table S5.** Cox proportional hazard model to assess risk factors associated with time to SARS-CoV-2 clearance adjusted for SARS-CoV-2 variant wave among adults and children in the SEARCh and OutSMART cohorts combined.

| **Risk Factors** | **Mean (SD)** | **HR (95% CI, *p*-value)** | **aHR (95% CI, *p*-value)** |
| --- | --- | --- | --- |
| Anti-Spike SIgA (SC-Ig)* | 1.6 (1.1) | 1.14 (1.05-1.25, p=0.003) | 1.51 (1.33-1.71, **p<0.001**) |
| Anti-Spike IgG* | 3.0 (1.2) | 0.92 (0.85-0.99, p=0.029) | 0.58 (0.49-0.69, **p<0.001**) |
| **COVID-19 vaccination** | **n (%)** |  |  |
| Not vaccinated | 782 (80.0) | (Reference) | (Reference) |
| Vaccinated | 195 (20.0) | 1.72 (1.39-2.12, p<0.001) | 1.30 (0.66-2.55, p=0.443) |
| **SARS-CoV-2 Variant** | **n (%)** |  |  |
| Wildtype | 481 (49.2) | (Reference) | (Reference) |
| Alpha | 328 (33.6) | 1.03 (0.87-1.22, p=0.714) | 1.13 (0.89-1.44, p=0.323) |
| Delta | 138 (14.1) | 1.68 (1.29-2.18, p<0.001) | 2.43 (1.15-5.14, **p=0.020**) |
| Omicron | 30 (3.1) | 2.13 (1.09-4.17, p=0.027) | 4.20 (1.70-10.37, **p=0.002**) |
| **Sex** | **n (%)** |  |  |
| Female | 501 (51.3) | (Reference) | (Reference) |
| Male | 476 (48.7) | 0.91 (0.78-1.05, p=0.202) | 1.05 (0.87-1.28, p=0.593) |
| **Age Group** |  |  |  |
| >=18 years | 711 (72.8) | (Reference) | (Reference) |
| 5-17 years | 90 (9.2) | 0.95 (0.74-1.21, p=0.661) | 1.22 (0.89-1.68, p=0.214) |
| <5 years | 176 (18.0) | 0.88 (0.73-1.06, p=0.194) | 1.00 (0.79-1.26, p=0.977) |
| **Race / Ethnicity** | **n (%)** |  |  |
| White, non-Hispanic | 585 (59.9) | (Reference) | (Reference) |
| Black, non-Hispanic | 199 (20.4) | 1.07 (0.86-1.31, p=0.555) | 0.75 (0.47-1.18, p=0.211) |
| Asian, non-Hispanic | 30 (3.1) | 0.75 (0.51-1.12, p=0.159) | 0.76 (0.54-1.08, p=0.127) |
| Multiracial, non-Hispanic | 73 (7.5) | 1.29 (0.96-1.74, p=0.091) | 1.28 (0.93-1.75, p=0.126) |
| Hispanic | 90 (9.2) | 1.12 (0.87-1.44, p=0.391) | 0.74 (0.50-1.10, p=0.134) |

**Note.** aHR = hazard ratio estimated from Cox proportional hazard model adjusted for all covariates presented in the table and clustered at the level of the individual participant. *log_10_ median fluorescence intensity (MFI). SC = secretory component. HR > 1.0 indicates higher probability of (and shorter time to) and HR < 1.0 indicates lower probability of (and longer time to) SARS-CoV-2 clearance. CI=confidence interval.

**Table S6.** Cox proportional hazard model to assess associations between time to anti-spike IgG seroconversion in oral fluid and time to SARS-CoV-2 clearance among unvaccinated adults in the SEARCh and OutSMART cohorts.

| **Risk Factors** | **Mean (SD)** | **HR (95% CI, *p*-value)** | **aHR (95% CI, *p*-value)** |
| --- | --- | --- | --- |
| Time to oral fluid anti-S IgG seroconversion (days) | 12.6 (12.9) | 0.98 (0.97-0.98, p<0.001) | 0.97 (0.96-0.98, p<0.001) |
| **Sex** | **n (%)** |  |  |
| Female | 277 (53.3) | (Reference) | (Reference) |
| Male | 243 (46.7) | 1.01 (0.82-1.25, p=0.91) | 1.02 (0.81-1.28, p=0.88) |
| **Age** |  |  |  |
| Up to 65 years | 461 (88.7) | (Reference) | (Reference) |
| Over 65 years | 59 (11.3) | 0.70 (0.48-1.01, p=0.06) | 1.09 (0.69-1.73, p=0.71) |
| **Race / Ethnicity** | **n (%)** |  |  |
| White, non-Hispanic | 270 (51.9) | (Reference) | (Reference) |
| Black, non-Hispanic | 138 (26.5) | 0.98 (0.74-1.28, p=0.87) | 1.36 (1.04-1.76, p=0.02) |
| Asian, non-Hispanic | 20 (3.8) | 0.67 (0.40-1.13, p=0.13) | 0.83 (0.65-1.06, p=0.12) |
| Multiracial, non-Hispanic | 25 (4.8) | 1.11 (0.62-1.99, p=0.73) | 1.33 (0.75-2.37, p=0.32) |
| Hispanic | 67 (12.9) | 1.03 (0.75-1.39, p=0.87) | 1.59 (0.99-2.56, p=0.054) |

**Note.** aHR = hazard ratio estimated from Cox proportional hazard model adjusted for all covariates presented in the table and clustered at the level of the individual participant. HR > 1.0 indicates higher probability of (and shorter time to) and HR < 1.0 indicates lower probability of (and longer time to) SARS-CoV-2 clearance. CI=confidence interval.

**Table S7.** Risk factors associated with time to COVID-19 symptoms resolution in children under 5 years in the SEARCh cohort.

| **Risk Factors** | **Mean (SD)** | **HR (95% CI, *p*-value)** | **aHR (95% CI, *p*-value)** |
| --- | --- | --- | --- |
| Anti-Spike SIgA (SC-Ig) * | 1.1 (1.1) | 1.08 (0.91-1.27, p<0.376) | 1.37 (1.16-1.61, **p<0.001**) |
| Anti-Spike IgG* | 2.5 (1.1) | 0.76 (0.64-0.90, p<0.002) | 0.52 (0.37-0.74, **p<0.001**) |
| **Gender** | **n (%)** |  |  |
| Female | 68 (37.6) | (Reference) | (Reference) |
| Male | 113 (62.4) | 1.12 (0.82-1.53, p<0.486) | 0.87 (0.60-1.27, p<0.471) |

**Note.** aHR = hazard ratio estimated from Cox proportional hazard model adjusted for all covariates presented in the table and clustered at the level of the individual participant. *log_10_ median fluorescence intensity (MFI). SC = secretory component. HR > 1.0 indicates higher probability of resolution of COVID-19 symptoms (and shorter symptoms duration) and HR < 1.0 indicates lower probability of resolution of COVID-19 symptoms (and longer symptoms duration). CI=confidence interval. Race/Ethnicity excluded due to small sample size in all but one subgroup.

**Table S8.** Risk factors associated with time to COVID-19 symptoms resolution in unvaccinated adults in the SEARCh and OutSMART cohorts.

| **Risk Factors** | **Mean (SD)** | **HR (95% CI, *p*-value)** | **aHR (95% CI, *p*-value)** |
| --- | --- | --- | --- |
| Anti-Spike SIgA (SC-Ig) * | 1.9 (1.1) | 1.16 (1.02-1.32, p<0.025) | 1.44 (1.26-1.65, **p<0.001**) |
| Anti-Spike IgG* | 2.8 (1.3) | 0.70 (0.64-0.78, p<0.001) | 0.53 (0.46-0.63, **p<0.001**) |
| **Gender** | **n (%)** |  |  |
| Female | 412 (55.5) | (Reference) | (Reference) |
| Male | 331 (44.5) | 1.18 (0.96-1.44, p<0.116) | 1.11 (0.86-1.43, p<0.408) |
| **Race / Ethnicity** | **n (%)** |  |  |
| White, non-Hispanic | 369 (49.7) | (Reference) | (Reference) |
| Black, non-Hispanic | 227 (30.6) | 0.96 (0.75-1.22, p<0.724) | 0.63 (0.41-0.95, **p<0.028**) |
| Asian, non-Hispanic | 20 (2.7) | 1.02 (0.64-1.61, p<0.943) | 0.79 (0.57-1.09, p<0.154) |
| Multiracial, non-Hispanic | 37 (5.0) | 2.05 (1.35-3.12, p<0.001) | 1.28 (0.58-2.81, p<0.535) |
| Hispanic | 90 (12.1) | 0.64 (0.45-0.91, p<0.012) | 0.60 (0.34-1.05, p<0.074) |

**Note.** aHR = hazard ratio estimated from Cox proportional hazard model adjusted for all covariates presented in the table and clustered at the level of the individual participant. *log_10_ median fluorescence intensity (MFI). SC = secretory component. HR > 1.0 indicates higher probability of resolution of COVID-19 symptoms (and shorter symptoms duration) and HR < 1.0 indicates lower probability of resolution of COVID-19 symptoms (and longer symptoms duration). CI=confidence interval.

**Table S9.** Association between oral fluid anti-Spike-RBD IgA and secretory Ig responses with time to SARS-CoV-2 clearance in days among children and adults in the SEARCh and OutSMART cohorts estimated using linear regression models with generalized estimating equations (GEE).

|  | **Spike (Mt. Sinai)** | | **RBD (Sino Biological)** | | **RBD (Mt. Sinai)** | | | |
| --- | --- | --- | --- | --- | --- | --- | --- | --- |
| **Oral fluid IgA** | **Beta (SE)** | ***p*-value** | **Beta (SE)** | ***p*-value** | **Beta (SE)** | | ***p*-value** | |
| IgA* | -3.4 (1.3) | **0.01** | -3.3 (1.6) | **<0.05** | -3.1 (1.6) | | **<0.05** | |
| IgG* | 0.9 (0.9) | 0.29 | 0.3 (0.9) | 0.74 | -0.3 (0.9) | | 0.76 | |
| **COVID-19 vaccination** |  |  |  |  |  | |  | |
| Unvaccinated | Reference |  | Reference |  | Reference | |  | |
| Vaccinated | 0.7 (2.6) | 0.79 | 0.7 (2.6) | 0.78 | 1.4 (2.6) | | 0.59 | |
| Weeks since symptoms onset | 0.3 (0.2) | 0.24 | 0.4 (0.2) | 0.12 | 0.4 (0.2) | | 0.05 | |
| **Age Group** |  |  |  |  |  | |  | |
| >=18 years | Reference |  | Reference |  | Reference | |  | |
| 5-17 years | -8 (3.2) | **0.01** | -7.6 (3.1) | **0.01** | -7.6 (3.1) | | **0.01** | |
| <5 years | -6.8 (2.7) | **0.01** | -7 (2.7) | **0.01** | -7.1 (2.6) | | **0.01** | |
| **Race / Ethnicity** |  |  |  |  |  | |  | |
| White, non-Hispanic | Reference |  | Reference |  | Reference | |  | |
| Black, non-Hispanic | 16.3 (6.7) | **0.02** | 16.4 (6.7) | **0.01** | 16.6 (6.9) | | **0.02** | |
| Asian, non-Hispanic | 6.7 (5.4) | 0.21 | 6.4 (6) | 0.28 | 5.9 (5.4) | | 0.27 | |
| Multiracial, non-Hispanic | 0.1 (2.2) | 0.97 | 0.6 (2.1) | 0.77 | 0.3 (2.2) | | 0.88 | |
| Hispanic | 0.1 (4.6) | 0.98 | -0.7 (5) | 0.88 | -0.8 (5) | | 0.87 | |
|  | **Spike (Mt. Sinai)** | | **RBD (Sino Biological)** | | **RBD (Mt. Sinai)** | | | |
| **Oral fluid secretory Ig** | **Beta (SE)** | ***p*-value** | **Beta (SE)** | ***p*-value** | **Beta (SE)** | | | ***p*-value** |
| SC Ig* (SIgA) | -2.9 (1.2) | **0.01** | -2.7 (1.3) | **0.04** | -2.3 (1.3) | | | 0.07 |
| IgG* | 0.6 (0.9) | 0.47 | -0.1 (0.8) | 0.94 | -0.8 (0.8) | | | 0.37 |
| Weeks since symptoms onset | 0.3 (0.2) | 0.25 | 0.4 (0.3) | 0.13 | 0.5 (0.2) | | | 0.05 |
| **COVID-19 vaccination** |  |  |  |  |  | | |  |
| Unvaccinated | Reference |  | Reference |  | Reference | | |  |
| Vaccinated | 0.5 (2.6) | 0.84 | 0.6 (2.6) | 0.81 | 1.5 (2.6) | | | 0.56 |
| **Age Group** |  |  |  |  |  | | |  |
| >=18 years | Reference |  | Reference |  | Reference | | |  |
| 5-17 years | -7.8 (3.1) | **0.01** | -7.4 (3) | **0.01** | -7.3 (3) | | | **0.02** |
| <5 years | -7.0 (2.6) | **0.01** | -7.1 (2.7) | **0.01** | -7.1 (2.6) | | | **0.01** |
| **Race / Ethnicity** |  |  |  |  |  | | |  |
| White, non-Hispanic | Reference |  | Reference |  | Reference | | |  |
| Black, non-Hispanic | 16.6 (6.9) | **0.02** | 16.5 (6.9) | **0.02** | 16.4 (7) | | | **0.02** |
| Asian, non-Hispanic | 6.4 (5.6) | 0.25 | 5.9 (5.9) | 0.32 | 5.7 (5.6) | | | 0.31 |
| Multiracial, non-Hispanic | 0.3 (2.2) | 0.88 | 0.5 (2.1) | 0.81 | 0.3 (2.2) | | | 0.89 |
| Hispanic | 0.1 (4.5) | 0.97 | -0.8 (4.7) | 0.87 | -1.1 (4.7) | | | 0.82 |

Note: GEE models with repeated measures (longitudinal oral fluid antibody data measurements) were adjusted for host factors and at the individual level. *log_10_ median fluorescence intensity (MFI). SC = secretory component.

**Table S10.** Association between oral fluid anti-Spike-RBD IgA and secretory Ig responses with COVID-19 symptoms duration in days among children and adults in the SEARCh and OutSMART cohorts estimated using linear regression models with generalized estimating equations (GEE).

|  | **Spike (Mt. Sinai)** | | | **RBD (Sino Biological)** | | | **RBD (Mt. Sinai)** | |
| --- | --- | --- | --- | --- | --- | --- | --- | --- |
| **Oral fluid IgA** | **Beta (SE)** | ***p*-value** | **Beta (SE)** | | ***p*-value** | **Beta (SE)** | | ***p*-value** |
| IgA* | -3.4 (1.6) | **0.04** | -4.9 (1.8) | | **<0.01** | -3.4 (1.7) | | **<0.05** |
| IgG* | 3.0 (1.6) | **0.03** | 3.8 (1.6) | | **0.01** | 2 (1.5) | | 0.17 |
| Weeks since symptoms onset | -0.1 (0.2) | 0.58 | -0.1 (0.3) | | 0.67 | 0.0 (0.3) | | 0.99 |
| **COVID-19 vaccination** |  |  |  | |  |  | |  |
| Unvaccinated | Reference |  | Reference | |  | Reference | |  |
| Vaccinated | 3.3 (4.3) | 0.43 | 2.8 (4.2) | | 0.50 | 3.3 (4.5) | | 0.46 |
| **Gender** |  |  |  | |  |  | |  |
| Female | Reference |  | Reference | |  | Reference | |  |
| Male | -3.6 (2.6) | 0.16 | -3.8 (2.5) | | 0.12 | -3.9 (2.6) | | 0.13 |
| **Age Group** |  |  |  | |  |  | |  |
| >=18 years | Reference |  | Reference | |  | Reference | |  |
| 0-17 years | -16.5 (2.7) | **0.01** | -16.3 (2.7) | | **<0.001** | -17.2 (2.8) | | **<0.001** |
| **Race / Ethnicity** |  |  |  | |  |  | |  |
| White, non-Hispanic | Reference |  |  | |  | Reference | |  |
| Black, non-Hispanic | 15.3 (5.6) | **<0.01** | 14.9 (5.3) | | **<0.01** | 15.8 (5.8) | | **<0.01** |
| Asian, non-Hispanic | -7.6 (6.4) | 0.23 | -7.2 (6.1) | | 0.24 | -8.9 (6.6) | | 0.18 |
| Multiracial, non-Hispanic | 4 (3.4) | 0.24 | 5.5 (3.4) | | 0.10 | 4.8 (3.3) | | 0.14 |
| Hispanic | 11.4 (6.6) | 0.08 | 11.1 (6.9) | | 0.11 | 10.6 (6.9) | | 0.12 |

|  | **Spike (Mt. Sinai)** | | | **RBD (Sino Biological)** | | | **RBD (Mt. Sinai)** | |
| --- | --- | --- | --- | --- | --- | --- | --- | --- |
| **Oral fluid secretory Ig** | **Beta (SE)** | ***p*-value** | **Beta (SE)** | | ***p*-value** | **Beta (SE)** | | ***p*-value** |
| SC Ig* (SIgA) | -3.3 (1.7) | 0.05 | -4.9 (1.9) | | **0.01** | -3.7 (1.8) | | **0.04** |
| IgG* | 2.9 (1.4) | **0.04** | 3.7 (1.6) | | **0.02** | 2.1 (1.5) | | 0.16 |
| Weeks since symptoms onset | -0.2 (0.3) | 0.54 | -0.1 (0.3) | | 0.63 | 0.0 (0.3) | | 0.90 |
| **COVID-19 vaccination** |  |  |  | |  |  | |  |
| Unvaccinated | Reference |  |  | |  | Reference | |  |
| Vaccinated | 2.9 (4.3) | 0.84 | 2.3 (4.3) | | 0.60 | 2.7 (4.6) | | 0.56 |
| **Gender** |  |  |  | |  |  | |  |
| Female | Reference |  |  | |  | Reference | |  |
| Male | -3.5 (2.6) | 0.16 | -3.9 (2.5) | | 0.12 | -4.1 (2.6) | | 0.12 |
| **Age Group** |  |  |  | |  |  | |  |
| >=18 years | Reference |  |  | |  | Reference | |  |
| 0-17 years | -16.8 (2.8) | **<0.001** | -16.5 (2.7) | | **<0.001** | -17.6 (2.9) | | **<0.001** |
| **Race / Ethnicity** |  |  |  | |  |  | |  |
| White, non-Hispanic | Reference |  |  | |  | Reference | |  |
| Black, non-Hispanic | 15.7 (5.7) | **<0.01** | 15.2 (5.4) | | **<0.01** | 15.9 (5.8) | | **<0.01** |
| Asian, non-Hispanic | -7.8 (6.3) | 0.22 | -7.7 (6.1) | | 0.21 | -8.9 (6.6) | | 0.18 |
| Multiracial, non-Hispanic | 4.3 (3.3) | 0.19 | 5.7 (3.3) | | 0.09 | 5.2 (3.3) | | 0.12 |
| Hispanic | 11.3 (6.7) | 0.09 | 10.8 (6.9) | | 0.11 | 10.4 (6.7) | | 0.12 |

Note: GEE models with repeated measures (longitudinal oral fluid antibody data measurements) were adjusted for host factors and at the individual level. *log_10_ median fluorescence intensity (MFI). SC = secretory component.

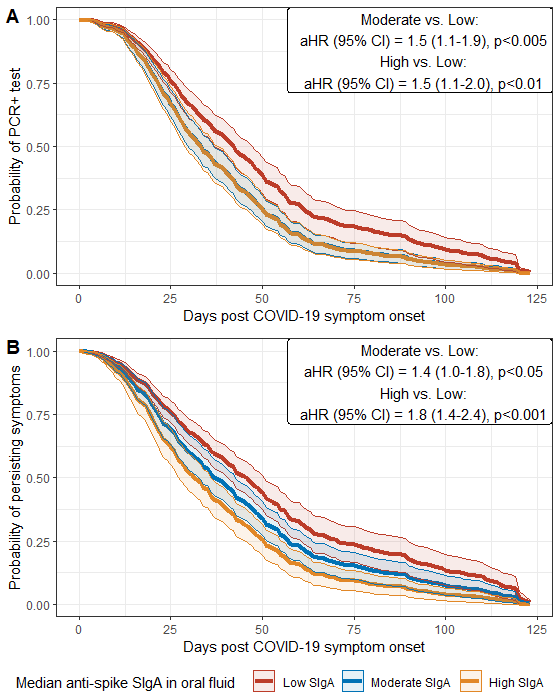

**Figure S1.** Kaplan-Meier survival curves stratified by each participant’s average oral fluid anti-spike SIgA response post COVID-19 symptoms onset (low; moderate and high tertile). Top: Probability of testing SARS-CoV-2 PCR positive over time in days. Bottom: Probability of persisting COVID-19 symptoms over time in days. aHR = hazard ratio estimated from Cox proportional hazard model adjusted for anti-spike IgG, COVID-19 vaccination status, age group, sex, and race and ethnicity, and clustered at the level of the individual participant.CI = confidence interval.

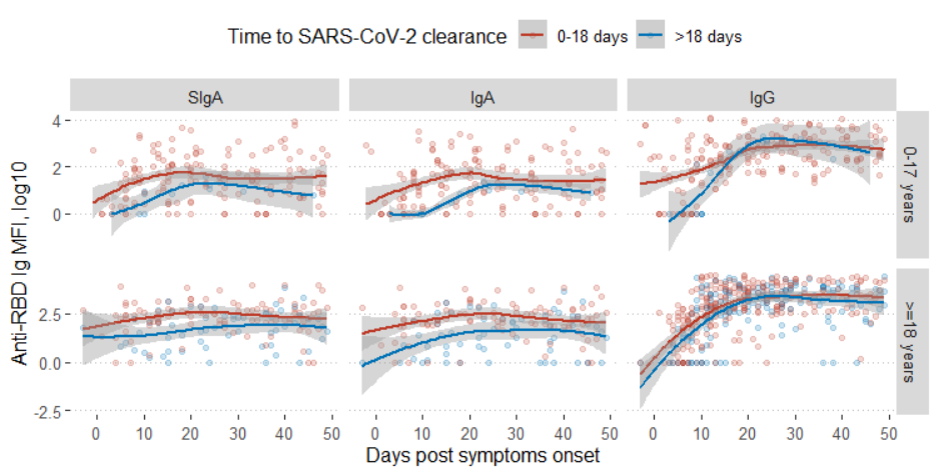

**Figure S2**. Anti-RBD antibody median fluorescence intensity (MFI) after COVID-19 symptoms onset stratified by time to SARS-CoV-2 clearance (time to testing SARS-CoV-2 PCR negative) and by age group.

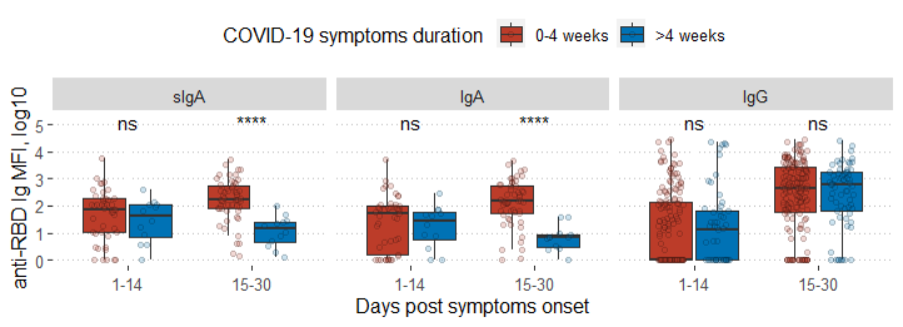

**Figure S3.** Oral fluid anti-SARS-CoV-2 RBD antibody up to 14 days and 15-30 days post COVID-19 symptoms onset stratified by COVID-19 symptoms duration in unvaccinated adults in the SEARCh and OutSMART study cohorts.

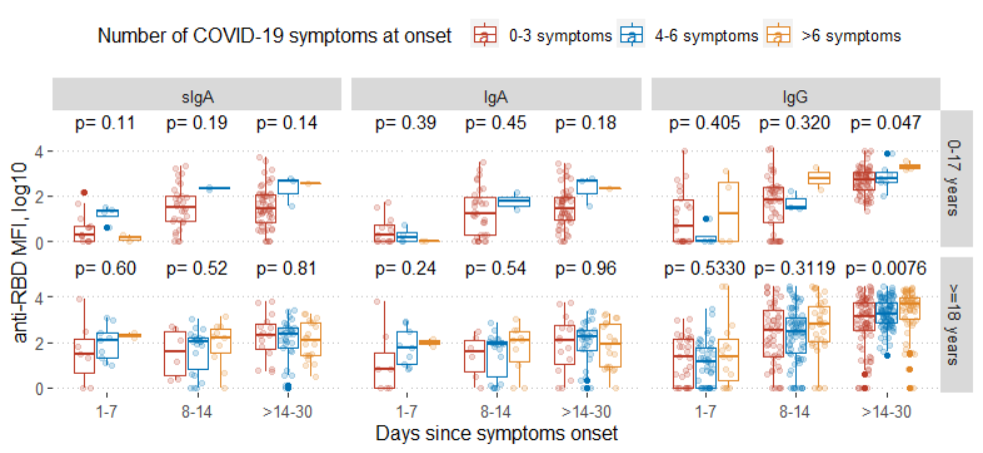

**Figure S4.** Oral fluid anti-SARS-CoV-2 RBD antibody levels stratified by the maximum number of COVID-19 symptoms reported during the illness period in adults and children in the SEARCh and OutSMART study cohorts.
